## Supplemental figures for "Improving SARS-CoV-2 variants monitoring in the absence of genomic surveillance capabilities: a serological study in Bolivian blood donors in October 2021 and June 2022"

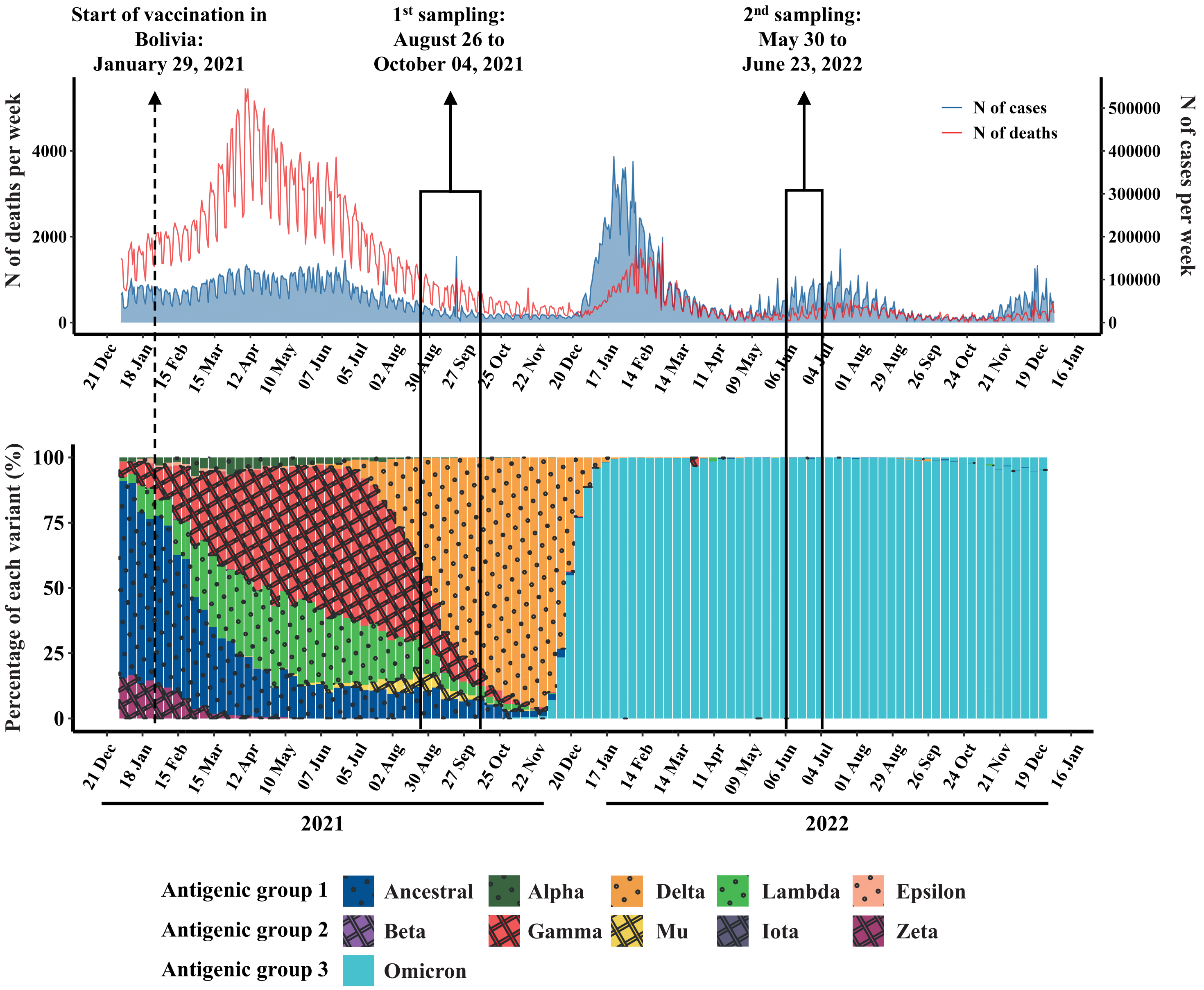


**Supplementary Figure S1: SARS-CoV-2 associated morbidity and mortality, and variant circulation in Bolivian surrounding countries (Argentina, Brazil, Chile, and Peru) in 2021 and 2022.**

Upper panel: Weekly number of cases (blue curve) and deaths (red curve) reported by the World Health Organization (WHO), were depicted.

Lower panel: Weekly variant prevalence obtained through national genomic surveillance and reported by the World Health Organization (WHO), were depicted.

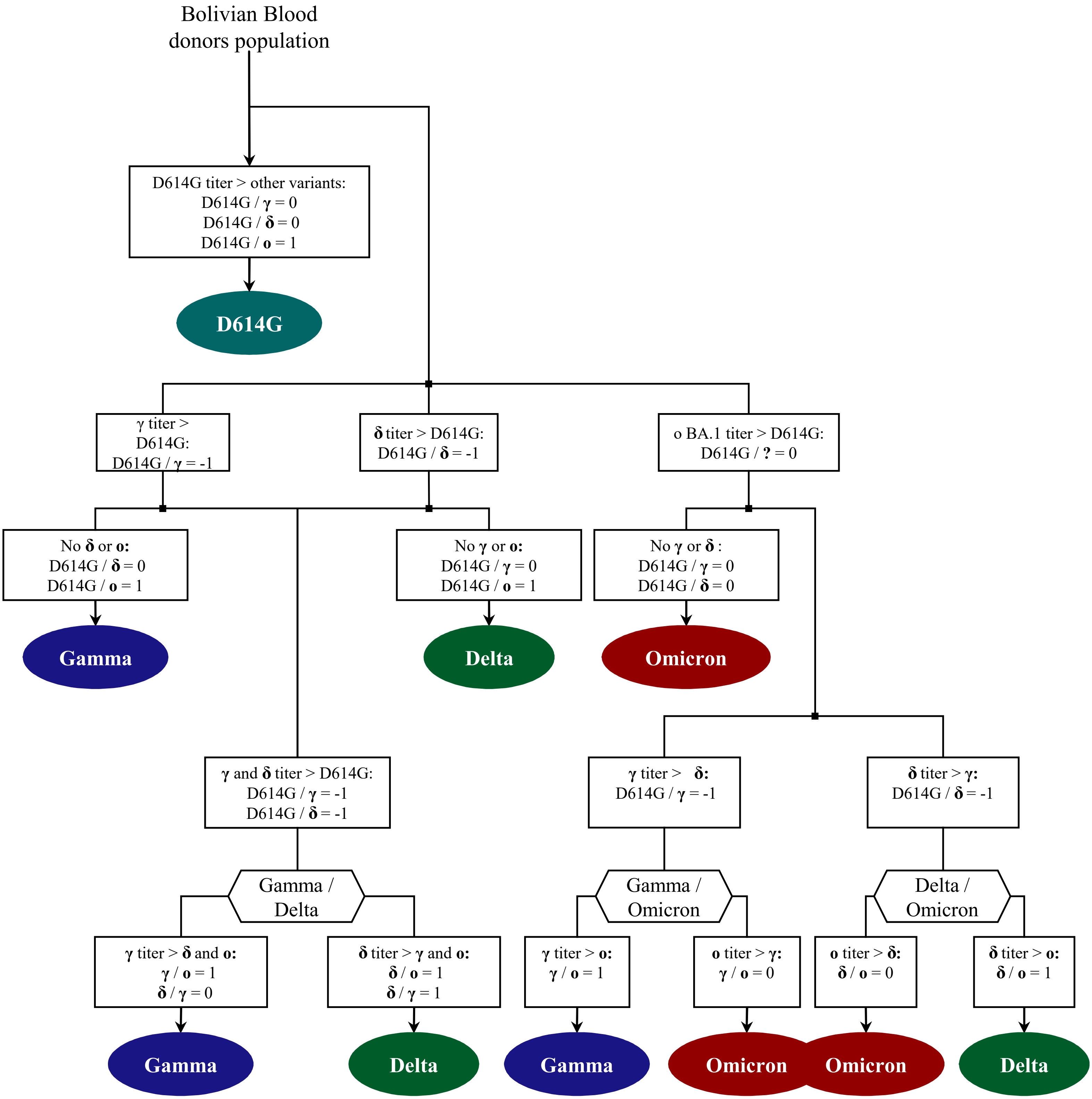


**Supplementary Figure S2: Flowchart for the identification of circulating variants.**
The base 2 logarithm of VNT titer ratios (log2-ratios) for all non-negative samples for the following variants pairs was calculated: D614G/Gamma (D614G/γ), D614G/Delta (D614G/δ), D614G/Omicron BA.1 (D614G/o), Delta/Gamma (δ/γ), Gamma/Omicron BA.1 (γ/o) and Delta/Omicron BA.1 (δ/o). Rules, established to determine which of the four variants tested had the highest VNT titer as described in supplementary methods, are summarized in the flowchart.

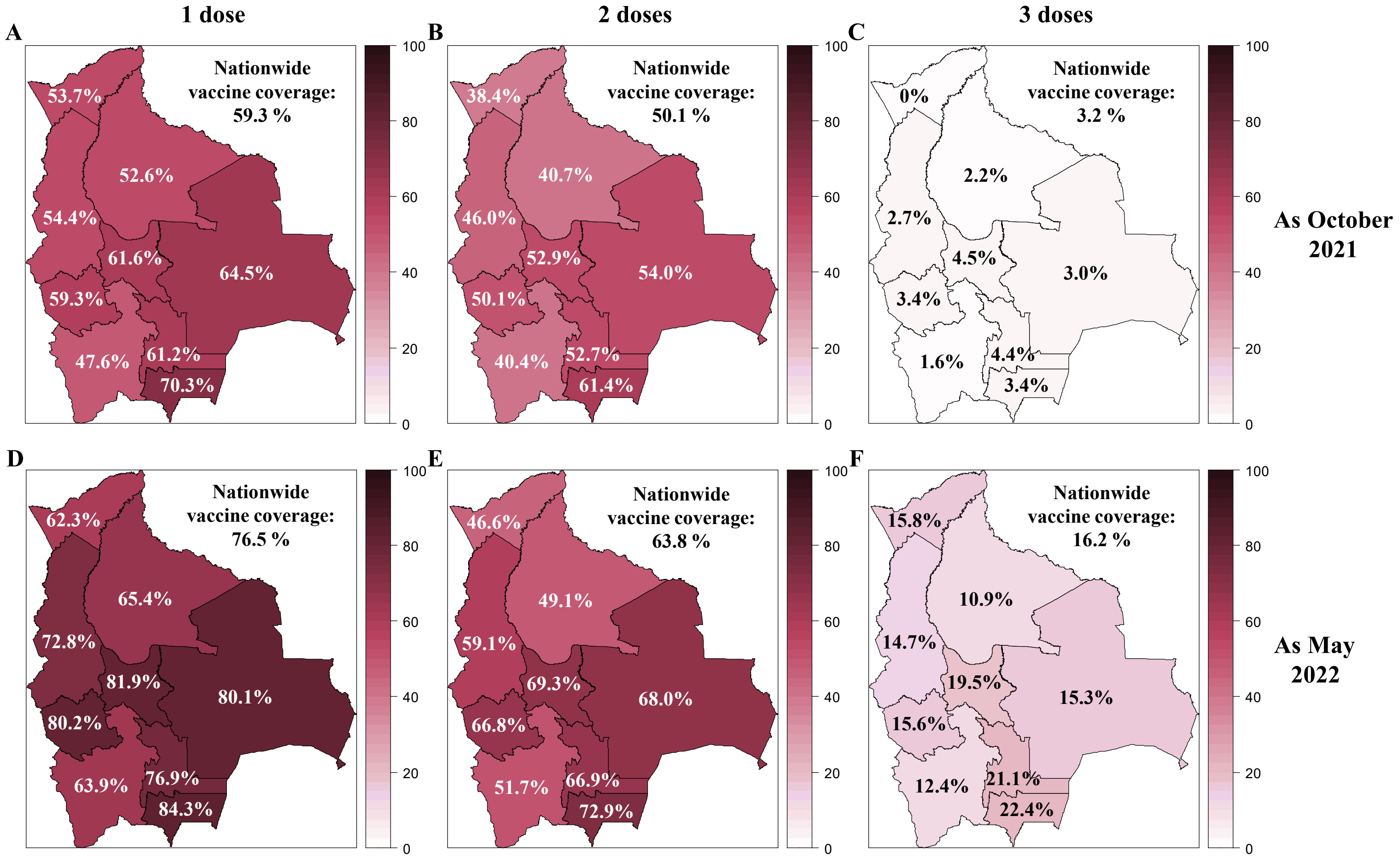


**Supplementary Figure S3: Vaccination coverage in Bolivia as October 2021 and May 2022.** The SARS-CoV-2 vaccine coverage for one dose (A, D), two doses (B, E), and three doses (C, F) was reported for each department in Bolivia and nationwide as October 2021 (A, B, C) and May 2022 (E, F, G). The Bolivian Ministry of Health has made public these data for adults over 18 years old in 2021 and people over 11 years old from 2022.

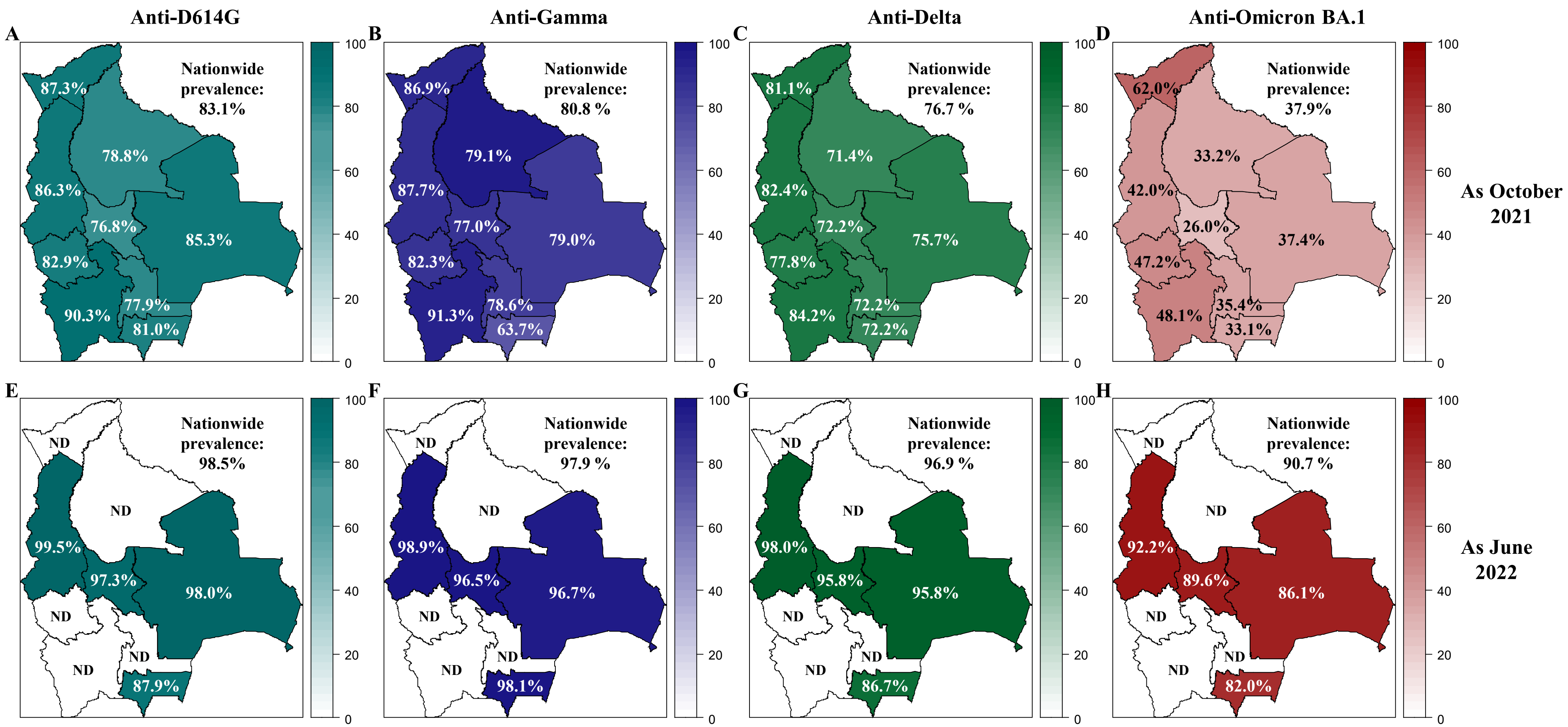


**Supplementary Figure S4: Prevalence of neutralizing anti-SARS-CoV-2 antibodies in Bolivia as October 2021 and June 2022.** The prevalence of neutralizing antibodies was reported for each department in Bolivia and nationwide for the D614G (A, E), Gamma (B, F), Delta (C, G) and Omicron BA.1 (D, H) variants as October 2021 (upper panel: A, B, C, D) and June 2022 (lower panel: E, F, G, H). The prevalence was obtained with the positivity cut-off ≥20.

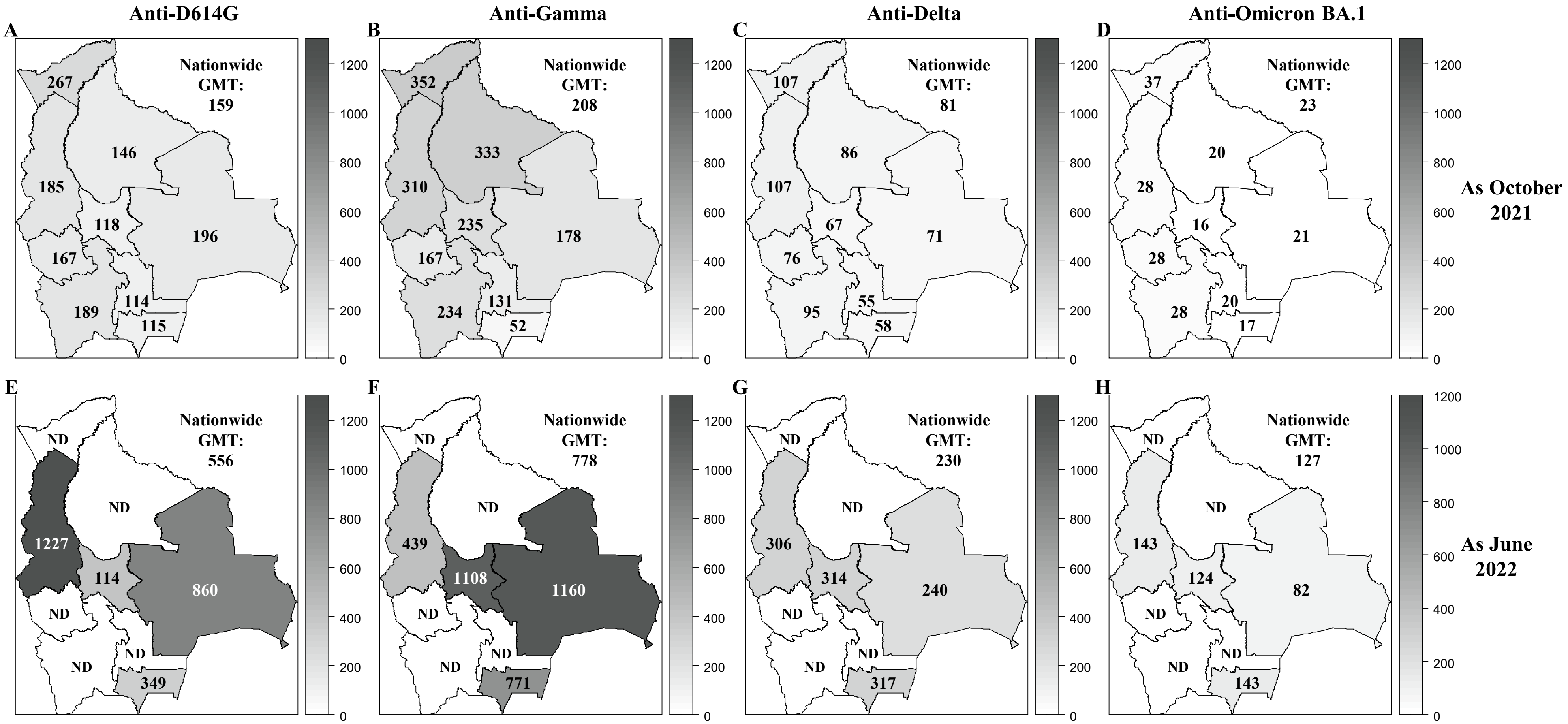


**Supplementary Figure S5: GMT titers of neutralizing anti-SARS-CoV-2 antibodies in Bolivia as October 2021 and June 2022.** The GMT titers of neutralizing antibodies was reported for each department in Bolivia and nationwide for the D614G (A, E), Gamma (B, F), Delta (C, G) and Omicron BA.1 (D, H) variants as October 2021 (upper panel: A, B, C, D) and June 2022 (lower panel: E, F, G, H).

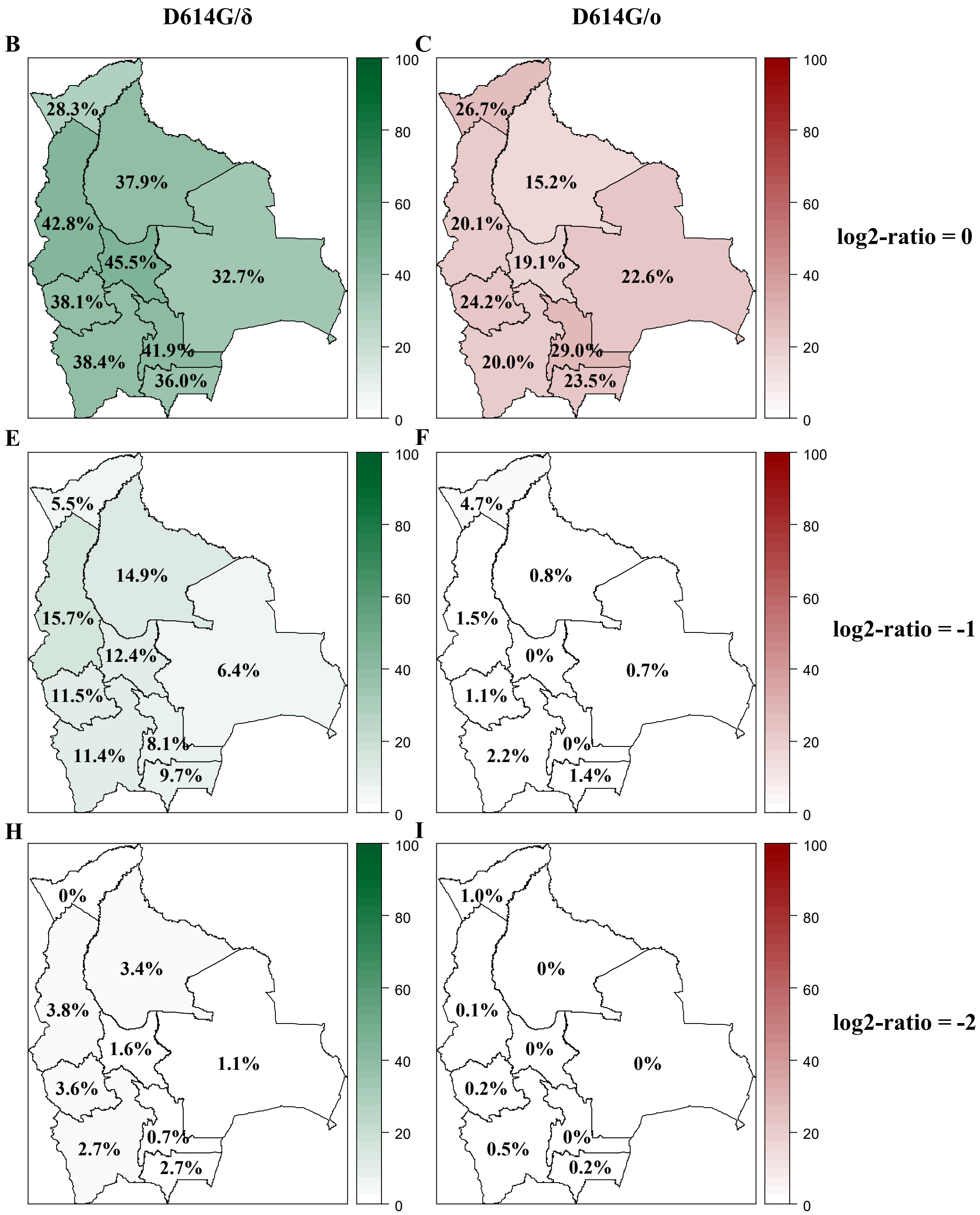


**Supplementary Figure S6: Variant circulation in Bolivia as October 2021.** Percentage of the population exhibiting titers for Gamma, Delta, and Omicron BA.1 being at least equivalent to (log2-ratios ≤0, upper panel: A, B), twice (log2-ratios ≤-1, middle panel: C, D), or four times (log2-ratios ≤-2, lower panel: E, F) that of D614G are presented for each department of Bolivia. Results obtained as October 2021 for D614G/δ and for D614G/o are depicted in green (A,C, E) and in red (B, D, F), respectively. D614G: ancestral D614G variant; γ: Gamma variant; δ: Delta variant; o: Omicron BA.1variant.

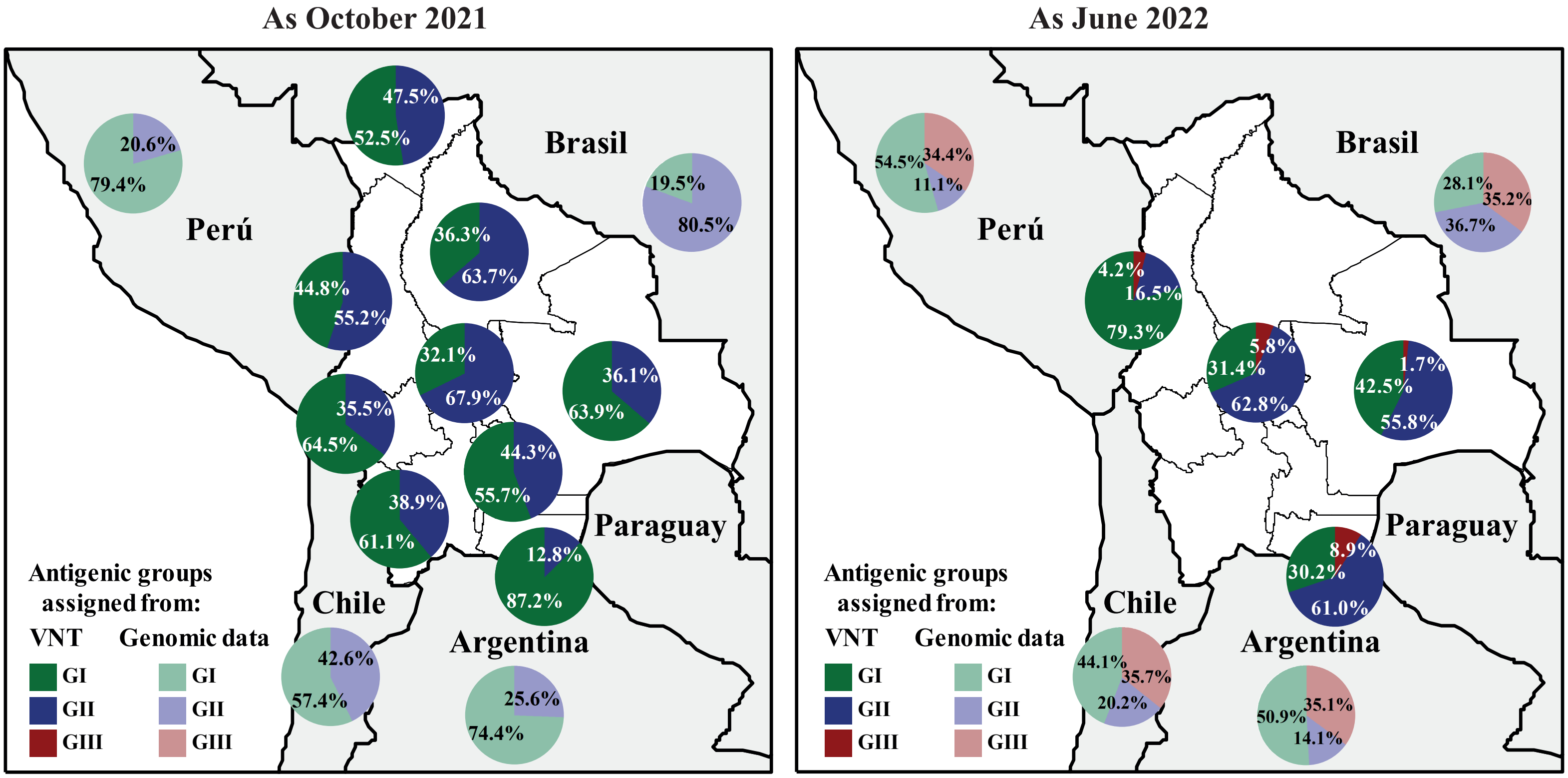


**Supplementary Figure S7: Quantification of SARS-CoV-2 circulating antigenic groups in Bolivia by Variant assignment method as October 2021 and as June 2022.**

Percentage of the population in each department presenting a neutralizing antibody response against a defined antigenic group, including the GI (green), GII (blue), GIII (red), using the Variant assignment method.

For antigenic group assignment by VNT, D614G and Delta variants were cluster in to the antigenic group I (GI), antigenic group II (GII) corresponded to the Gamma variant, while antigenic group III (GIII) was defined by the Omicron BA.1 variant (see methods for details). For evaluating genomic data, the prevalences of GI (from ancestral, Alpha, Delta, Lambda, and Epsilon variants; light-green), the prevalences of GII (from Beta, Gamma, Mu, Iota, and Zeta variants; light-blue), and the prevalences of GIII (from all Omicron-derived variants; light-red) were obtained for neighboring countries of Bolivia using publicly accessible genomic data when available.

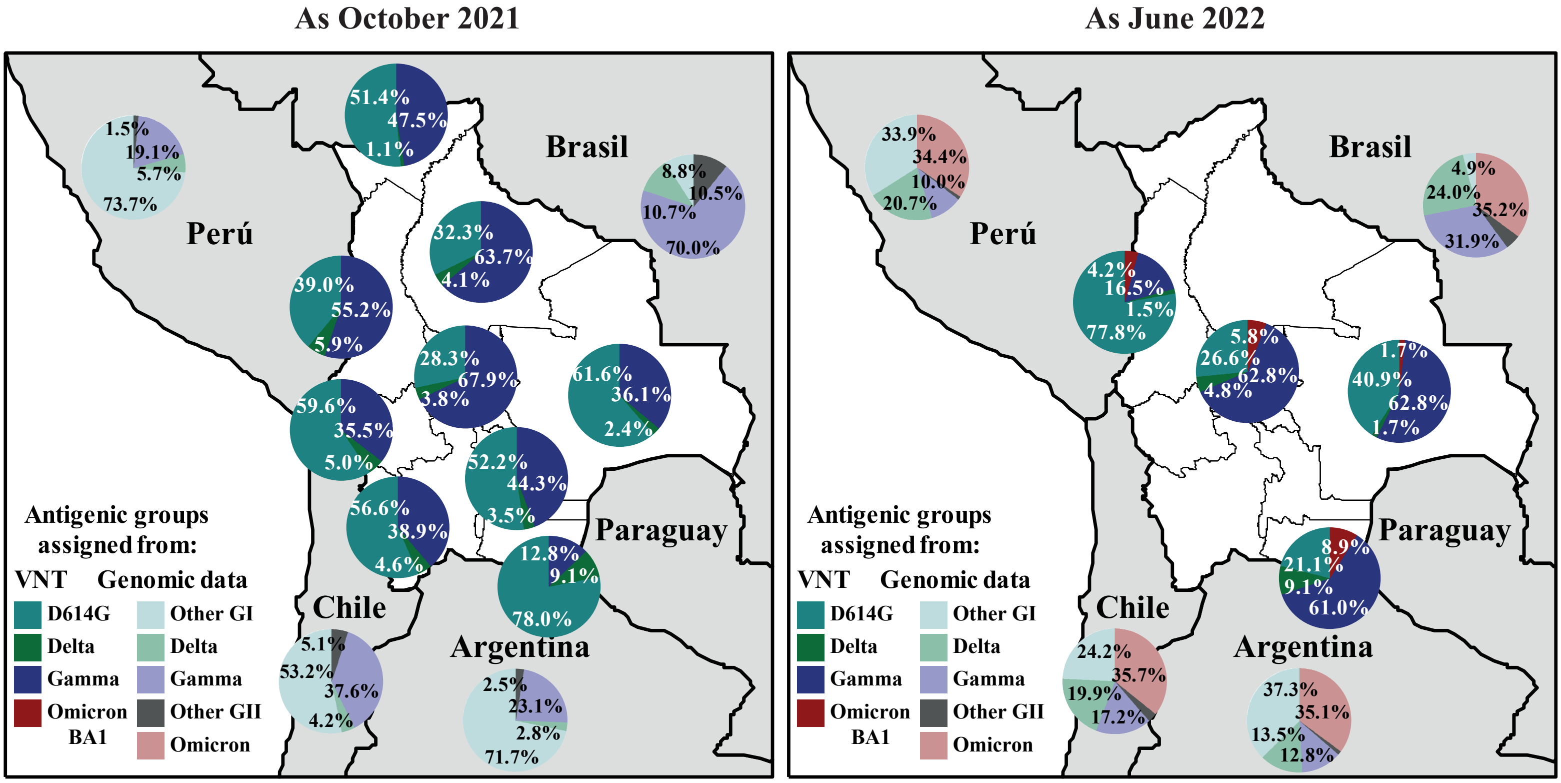


**Supplementary Figure S8: Quantification of SARS-CoV-2 circulating antigenic groups in Bolivia by Rules method as October 2021 and as June 2022.**

Percentage of the population in each department presenting a neutralizing antibody response against a defined variant, including D614G (cyan), Delta (green), Gamma (blue), and Omicron BA.1 (red), using the Rules method (see methods for details). For evaluating genomic data, the prevalences of ancestral, Alpha, Lambda, and Epsilon variants were grouped as Other GI (light-cyan), and the prevalences of Beta, Mu, Iota, and Zeta variants were grouped as Other GII (grey). The prevalences of all Omicron-derived variants were grouped as Omicron (light-red). Prevalence of Delta (light-green) and Gamma (light-blue) were also obtained for neighboring countries of Bolivia using publicly accessible genomic data when available.
