## Supplementary Table 1 for "Improving SARS-CoV-2 variants monitoring in the absence of genomic surveillance capabilities: a serological study in Bolivian blood donors in October 2021 and June 2022"

**Supplementary Table 1: SARS-CoV-2 hybrid immunity estimation by department in 2021 and 2022.**

The hybrid immunity was estimated based on the prevalence of anti-S1 and anti-NCP antibodies.

| Departments | 2021 | 2022 |
|---|---|---|
| Beni | 58.9% | ND |
| Cochabamba | 55.8% | 83.6% |
| La Paz | 57.8% | 89.8% |
| Oruro | 58.2% | ND |
| Pando | 51.4% | ND |
| Potosi | 73.6% | ND |
| Santa Cruz | 54.7% | 83.3% |
| Sucre | 67.7% | ND |
| Tarija | 60.2% | 90.6% |
